# Impact of the disposable vape ban in Great Britain: a representative interrupted time-series study 2022-2026

**DOI:** 10.64898/2026.06.09.26355241

**Authors:** Harry Tattan-Birch, Sarah E. Jackson, Vera Buss, Sharon Cox, Lion Shahab, Loren Kock, Linda Bauld, Jamie Brown

## Abstract

**Objective:** To examine changes in vaping and smoking trends following the announcement and implementation of the disposable vape ban in Great Britain.

**Design:** Interrupted time-series analysis of representative monthly cross-sectional data from the Smoking Toolkit Study.

**Setting:** Great Britain.

**Participants:** 118,946 adults (≥16y), including 12,042 young adults (16-24y), surveyed between Jan-2022 and Feb-2026.

**Main outcome measures:** Changes in trends in disposable vape use among vapers, and current vaping and smoking prevalence, using seasonally-adjusted generalised additive models with comparisons against a no-ban counterfactual in which pre-announcement trends continued unchanged.

**Results:** The proportion of vapers mainly using disposable devices began to decline following the announcement of the ban in Jan-2024, with the fall accelerating after implementation in June-2025. By Feb-2026, 5.6% (95%CI 4.6-6.9) of adult vapers and 7.1% (5.1-10.1) of young adult vapers mainly used disposables, compared with 62.0% (53.6-71.8) and 63.6% (52.7-76.7), respectively, under a no-ban counterfactual. Increases in vaping prevalence slowed post-announcement and plateaued post-implementation; by Feb-2026, prevalence was lower than the no-ban counterfactual in adults (13.6% v 18.8%; difference −5.2 percentage points, 95%CI −7.1 to −3.3) and young adults (27.8% v 39.1%; −11.3, −18.6 to −4.1). Declines in smoking prevalence stalled among adults and reversed among young adults post-announcement, before shifting downward again post-implementation; by Feb-2026, smoking prevalence was similar to the no-ban counterfactual in adults (difference +0.9 percentage points, −0.5 to +2.2) but possibly higher in young adults (+3.3, −0.5 to +7.1).

**Conclusions:** The disposable vape ban in Great Britain was associated with substantial changes after both announcement and implementation, including a marked reduction in disposable vape use and a slowing then plateauing of growth in overall vaping prevalence. However, declines in smoking also temporarily slowed—and among young adults, reversed—after the announcement, before downward trends resumed after implementation.

**What is already known on this topic:**

- A ban on disposable vapes across the UK was announced in January 2024 and implemented in June 2025, following a rapid rise in use, particularly among youth and young adults.
- Early data indicated a shift from disposable to reusable vapes following the announcement of the ban.
- However, evidence is limited on the impact of the ban’s actual implementation on disposable vape use and on broader changes in vaping and smoking prevalence.

**What this study adds:**

- Use of disposable vapes among people who vape declined after the announcement of the ban and fell more steeply after it was implemented.
- Growth in vaping prevalence slowed after the announcement and plateaued after implementation, both overall and in young adults.
- Declines in smoking prevalence slowed among all adults, and among young adults reversed after the announcement, before resuming after implementation.

## Introduction

The use of disposable (also known as “single-use”) nicotine vapes increased rapidly in Great Britain from Spring 2021, especially among young people.[1,2] By design, these products are thrown away once the battery or e-liquid runs out, despite containing lithium-ion batteries and other components that could, in principle, be reused or recycled.[3,4] While recycling facilities exist, these are not widespread nor easily accessible for many users, and only an estimated 17% of users recycle their devices.[5] The rise in popularity of disposable vapes was accompanied by notable increases in overall nicotine use in youth and young adults, and greater uptake of vaping among those who had never regularly smoked.[6–9] In response, the UK Government announced on 28 January 2024 its intention to ban the sale and supply of disposable vapes.[10] This legislation came into force on 1 June 2025 across all four nations of the UK.

Early analyses indicated that trends in device use began to shift after the announcement, even before the ban was implemented. The upward trend in the proportion of vapers mainly using disposable devices reversed, though it remained around 30% in January 2025.[11] Changes were most pronounced among younger adults (falling from 63% to 35%), the group in which disposable vape use had increased most rapidly before the announcement. However, evidence is lacking on whether such changes extended beyond vaping device type to influence smoking or overall nicotine use.

Moreover, there is currently no evidence on changes following implementation of the disposable vape ban in June 2025. Implementation may have led to a further reduction in the proportion of people vaping who use disposables. However, there are media reports of continued availability of disposable vapes in shops after the ban, suggesting that use may have continued.[12] Beyond changes in device type, the ban may also have influenced overall vaping prevalence, for example by potentially reducing the attractiveness or accessibility of vaping products, both overall and to young people specifically. It may also have had effects on smoking, either by reducing the use of vapes as a smoking cessation aid—given evidence that vaping during quit attempts increased following the rise in disposable vape use from 2021[13]—or, as suggested by qualitative interviews, by leading some individuals who have developed nicotine dependence to smoke cigarettes instead.[14]

Using representative monthly data from people aged ≥16 years in Great Britain up to February 2026, this study aimed to examine whether the January 2024 announcement and June 2025 implementation of the disposable vape ban were followed by changes in:

1. use of disposable vapes as the main device type among people who vape; and
2. current prevalence of (i) vaping, (ii) smoking, and (iii) any nicotine use in the overall population.

All outcomes are reported for all adults (aged ≥16 years) and separately for young adults (aged 16–24 years).

## Methods

### Pre-registration

The protocol was pre-registered on the Open Science Framework prior to data analysis (https://osf.io/beac8/).

### Data source

The Smoking Toolkit Study (STS) is a nationally representative, repeat cross-sectional survey that recruits approximately 2,450 adults (aged ≥16) in Great Britain (England, Wales, and Scotland) each month.[7,15] Samples are newly drawn each month, with data collection taking place during the middle weeks of the month, using a combination of random probability and quota sampling. Validation studies indicate that the survey yields nationally representative estimates, closely matching other national surveys that rely exclusively on random probability sampling across key variables (including sex, age, socioeconomic position, smoking status, and cigarette consumption) as well as data on tobacco sales.[16,17] Data are collected via telephone interviews, with one participant per household interviewed by a trained interviewer.

### Design

This study used an interrupted time-series design with two pre-specified interruption breakpoints, drawing on monthly repeat cross-sectional data from the STS. The analytical time series ran from January 2022 to February 2026. January 2022 was selected as the start point to align with the time period used in previous analyses of disposable vape use,(11) capturing the period during which disposable vapes became established in the market while providing sufficient pre-announcement observations.[11]

Changes in the underlying trend were permitted at two breakpoints. The first corresponds to the government’s announcement on 28th January 2024 of its intention to ban disposable vapes; February 2024 was treated as the first post-announcement month because the announcement occurred late in January, after that month’s STS data collection period had ended. The second breakpoint corresponds to implementation of the ban from 1st June 2025, with June 2025 treated as the first post-implementation month.

For descriptive purposes, the time series was characterised as covering a pre-announcement period (January 2022–January 2024), a post-announcement/pre-implementation period (February 2024–May 2025), and a post-implementation period (June 2025–February 2026).

### Participants

The sample included all participants (aged ≥16 years) who responded to the STS during the study period (January 2022 to February 2026, inclusive) and reported their current smoking and vaping status.

### Measures

#### Outcomes

***Current smoking*** was defined as current use of cigarettes (daily or non-daily) or any other combustible tobacco product (e.g., cigars or shisha).

***Current vaping*** was assessed through questions that asked about current use of a selection of nicotine and tobacco products. Interviewers read out the selection to each participant, prompting them to identify any products they were currently using. These questions included a general catch-all item asking about use for any reason, as well as (depending on smoking status) questions about use to cut down on smoking or in places where smoking is prohibited. Participants were classified as currently vaping if they reported using a “Electronic cigarette or vaping device” and/or “Juul” in response to any of these questions.

***Any current nicotine use*** was defined based on reporting use of any of the nicotine and tobacco products listed (including vapes, nicotine pouches,[18] heated tobacco products,[19] and nicotine replacement therapy) in response to above questions, as well as those who reported current smoking.

***Use of disposable vapes as the main device type*** was assessed among people who reported current vaping using the following question: “Which of the following do you mainly use?”. Response options were: (a) a disposable e-cigarette or vaping device; (b) an e-cigarette or vaping device that uses replaceable pre-filled cartridges (rechargeable); (c) an e-cigarette or vaping device with a tank that you refill with liquids (rechargeable); (d) a modular system that you refill with liquids (using your own combination of separate devices such as batteries and atomizers); or (e) don’t know. For analysis, device types were grouped into disposable (response a) and reusable (responses b–d). Participants who responded (e) were excluded from the main analytic sample. However, responses across all device categories, including “don’t know”, were presented descriptively.

#### Demographic and contextual variables

***Country of residence*** was recorded (England, Scotland, Wales) by interviewers.

***Age*** was categorised into six groups (16–17, 18–24, 25–34, 35–44, 45–64, and ≥65 years) for descriptive analyses. For the primary analyses, we examined use overall across all ages and in those aged 16–24 years, reflecting evidence that increases in disposable vape use were greatest in this age range[20] and that reducing vaping among young people was a central aim of the disposable vape ban.[10] This approach is consistent with our previous analysis examining changes following the announcement of the ban.(5)

***Gender*** identity was determined through the question, “Which of the following best describes how you think of yourself?” Responses were coded “Man”, “Woman”, or “In another way”.

***Social grade*** was determined using the National Readership Survey (NRS) social grade classification, based on the occupation of the chief income earner in the household. It was grouped into ABC1 (managerial, professional, and intermediate occupations, representing more advantaged groups) and C2DE (skilled manual, semi-skilled and unskilled manual occupations, and those not in employment, representing less advantaged groups).[7,21]

***Seasonality*** was modelled non-linearly using cyclic cubic splines of calendar month (details below).

### Statistical analysis

All analyses were conducted in R version 4.5.2. The STS uses raking (iterative proportional fitting) to construct survey weights that align the sample with the population of Great Britain.[15,17,22] All analyses were survey weighted, and participants with missing data on smoking or vaping were excluded. Estimates were presented with 95% confidence intervals (CIs).

We used segmented regression to assess changes in monthly trends in each outcome (later converted to annual trends for interpretability). Models were estimated using generalised additive models (GAMs) with a log link, allowing effects to be interpreted as proportional changes in prevalence over time (i.e., yearly risk ratios represent the annual relative percentage change in the outcome).

Two interruption breakpoints were modelled: the ban announcement (28 January 2024; February 2024 as the first post-announcement month) and ban implementation (1 June 2025; June 2025 as the first post-implementation month). Time was indexed as months since January 2022 (January 2022 = 0 to February 2026 = 49) and used to model the underlying secular trend.

Changes in trend were captured with two time-since terms: a post-announcement slope term (0 through January 2024, then increasing by 1 each month from February 2024) and a post-implementation slope term (0 through May 2025, then increasing by 1 each month from June 2025). As preregistered, models estimated changes in slope only and did not include step changes. Comparison of fitted values with observed monthly estimates showed no evidence of abrupt discontinuities at either breakpoint.[23]

We adjusted for seasonality using calendar month (ranging 1–12) modelled with a low-dimensional cyclic cubic spline (k=6), allowing for repeating month-of-year effects while avoiding overfitting.[24] For presentation, fitted trends were shown as seasonally-adjusted predictions by averaging predicted values over months 1–12 at each time point.

Analyses were conducted for all adults (aged ≥16 years) and separately for young adults (aged 16–24 years). The primary outcome was the proportion mainly using disposable devices among people who vape. Secondary outcomes were current prevalence of vaping, smoking, and any nicotine use, each estimated among all adults or young adults respectively. Each outcome was assessed in a separate model. Additional analyses relating to use of vapes in smoking quit attempts are reported separately because interpretation was complicated by overlap with the contemporaneous Swap to Stop programme introduced in December 2023.[25]

To aid interpretation, we reported both changes in trend at each breakpoint and model-based counterfactual contrasts (i.e., what prevalence may have been, under certain assumptions, if the disposable vape ban had not been announced or implemented). Counterfactual predictions were generated under a “no-ban” scenario in which the pre-announcement trend continued unchanged (i.e., both post-announcement and post-implementation slope-change terms set to zero). We compared these counterfactual predictions with the actual (modelled) predictions from the primary model, and expressed effects as absolute differences in seasonally-adjusted predicted prevalence in February 2026, the final month included in this study. Confidence intervals for modelled estimates and absolute differences were calculated from the model coefficient covariance matrix using the delta method.

We also conducted two supplementary analyses. First, we descriptively examined the distribution of responses to the main vaping device type question (including the “don’t know” category) among vapers across each quarter. Second, in an unplanned analysis, we repeated interrupted time-series models separately for daily and non-daily smoking prevalence.

### Patient and public involvement

The STS convenes a diverse patient and public involvement (PPI) group several times each year, comprising individuals with a range of nicotine use experiences. The research team also regularly engages with members of the public and stakeholders from the Department of Health and Social Care and Action on Smoking and Health through presentations and meetings.

Insights from these activities inform the study’s overall research priorities and specific research questions. In addition, interviewers routinely invite participant feedback on the questionnaire each month.

## Results

### Sample

A total of 119,724 participants aged ≥16 years were surveyed in Great Britain between January 2022 and February 2026. Of these, 118,946 (99.4%) provided data on their smoking and vaping status, and were thus included in the analytic sample (this included 12,042 young adults aged 16–24 years). The weighted sociodemographic profile of the sample was similar across pre-announcement, post-announcement/pre-implementation, and post-implementation periods (Table S1).

Analyses of the primary outcome, mainly using disposable vapes, were restricted to respondents who reported current vaping and provided information on main device type used (N=11,115 adults; N=2,473 young adults). Analyses of prevalence outcomes included the entire analytic sample.

### Use of disposable vapes

There were substantial changes in trends in the proportion of vapers mainly using disposable devices after both the announcement and implementation of the disposable vape ban (**Table 1**; **Figure 1A**). Among all adults, use of disposable vapes was increasing before the January 2024 announcement (yearly risk ratio [RR_trend_]=1.18, 95% CI=1.12–1.24) and had reached around 42%, but this trend reversed (change in yearly risk ratio RR_Δtrend_=0.57, 95% CI=0.51–0.63), corresponding to a post-announcement decline (RR_trend_=0.67, 95% CI=0.62–0.71). There was a further downward change in trend after implementation in June 2025 (RR_Δtrend_=0.20, 95% CI=0.14–0.28), leaving a steep post-implementation decline (RR_trend_=0.13, 95% CI=0.10–0.18). By February 2026, the observed prevalence of mainly using disposables among adult vapers was 5.6% (95% CI=4.6–6.9), compared with 62.0% (53.6–71.8) in the no-ban counterfactual scenario, an absolute difference of -56.4 percentage points (-65.5 to -47.3).

**Table 1.** Trends in disposable vape use and nicotine use before and after announcement (January 2022) and implementation (June 2025) of the disposable vape ban in Great Britain.

|  | Among current vapers <sup>a</sup> | Among all respondents <sup>b</sup> |  |  |
| --- | --- | --- | --- | --- |
|  | Mainly use disposables | Current vaping | Current smoking | Any current nicotine use |
| **All adults** |  |  |  |  |
| **Annual trends, RR [95% CI]** |  |  |  |  |
| Pre-announcement trend | 1.18 [1.12–1.24] | 1.20 [1.17–1.25] | 0.96 [0.93–0.98] | 1.04 [1.02–1.07] |
| Change in trend after announcement | 0.57 [0.51–0.63] | 0.91 [0.85–0.97] | 1.07 [1.01–1.14] | 1.03 [0.99–1.08] |
| Post-announcement trend | 0.67 [0.62–0.71] | 1.09 [1.04–1.14] | 1.03 [0.99–1.07] | 1.08 [1.05–1.11] |
| Change in trend after implementation | 0.20 [0.14–0.28] | 0.86 [0.75–0.98] | 0.89 [0.79–1.00] | 0.89 [0.81–0.97] |
| Post-implementation trend | 0.13 [0.10–0.18] | 0.94 [0.84–1.04] | 0.91 [0.83–1.00] | 0.96 [0.89–1.03] |
| **End-of-series prevalence (Feb 2026), % [95% CI]** |  |  |  |  |
| Observed | 5.6 [4.6–6.9] | 13.6 [12.8–14.4] | 15.2 [14.4–16.0] | 25.9 [24.9–26.9] |
| No-ban counterfactual scenario | 62.0 [53.6–71.8] | 18.8 [17.0–20.7] | 14.3 [13.2–15.5] | 26.4 [24.8–28.1] |
| Difference | -56.4 [-65.5 to -47.3] | -5.2 [-7.1 to -3.3] | 0.9 [-0.5 to 2.2] | -0.6 [-2.4 to 1.3] |
| **Young adults (16–24 years)** |  |  |  |  |
| **Annual trends, RR [95% CI]** |  |  |  |  |
| Pre-announcement trend | 1.01 [0.94–1.07] | 1.23 [1.16–1.31] | 0.92 [0.87–0.98] | 1.08 [1.04–1.13] |
| Change in trend after announcement | 0.59 [0.51–0.69] | 0.88 [0.78–0.99] | 1.22 [1.06–1.40] | 1.05 [0.96–1.15] |
| Post-announcement trend | 0.60 [0.54–0.66] | 1.08 [1.00–1.17] | 1.12 [1.02–1.23] | 1.14 [1.07–1.21] |
| Change in trend after implementation | 0.23 [0.13–0.41] | 0.91 [0.71–1.16] | 0.75 [0.56–1.00] | 0.85 [0.70–1.02] |
| Post-implementation trend | 0.14 [0.08–0.23] | 0.98 [0.81–1.18] | 0.84 [0.66–1.05] | 0.96 [0.84–1.11] |
| **End-of-series prevalence (Feb 2026), % [95% CI] <sup>c</sup>** |  |  |  |  |
| Observed | 7.1 [5.1–10.1] | 27.8 [25.0–30.8] | 19.5 [17.2–22.2] | 40.6 [37.5–43.9] |
| No-ban counterfactual scenario | 63.6 [52.7–76.7] | 39.1 [32.8–46.7] | 16.2 [13.4–19.6] | 41.5 [36.3–47.4] |
| Difference | -56.5 [-68.5 to -44.4] | -11.3 [-18.6 to -4.1] | 3.3 [-0.5 to 7.1] | -0.9 [-7.0 to 5.2] |
| **Abbreviations:** RR, risk ratio; CI, confidence interval. **Notes:** Results are derived from survey-weighted segmented regression analyses using generalised additive models with a log link, adjusted for seasonality. Table S2 shows specific prevalence estimates at key time points surrounding the announcement and implementation of the ban. |  |  |  |  |
| <sup>a</sup> Among respondents who reported current vaping and provided information on main device type used (N=11,115 adults; N=2,473 young adults aged 16–24). |  |  |  |  |
| <sup>b</sup> Among all respondents who provided their smoking and vaping status (N=118,946 adults; N=12,042 young adults aged 16–24). |  |  |  |  |
| <sup>c</sup> No-ban estimates represent model-based predictions at February 2026 obtained by extrapolating the pre-announcement trend forward. The observed–no ban difference is calculated as the modelled observed prevalence minus the extrapolated no-ban prevalence at February 2026. |  |  |  |  |

**Figure 1.**
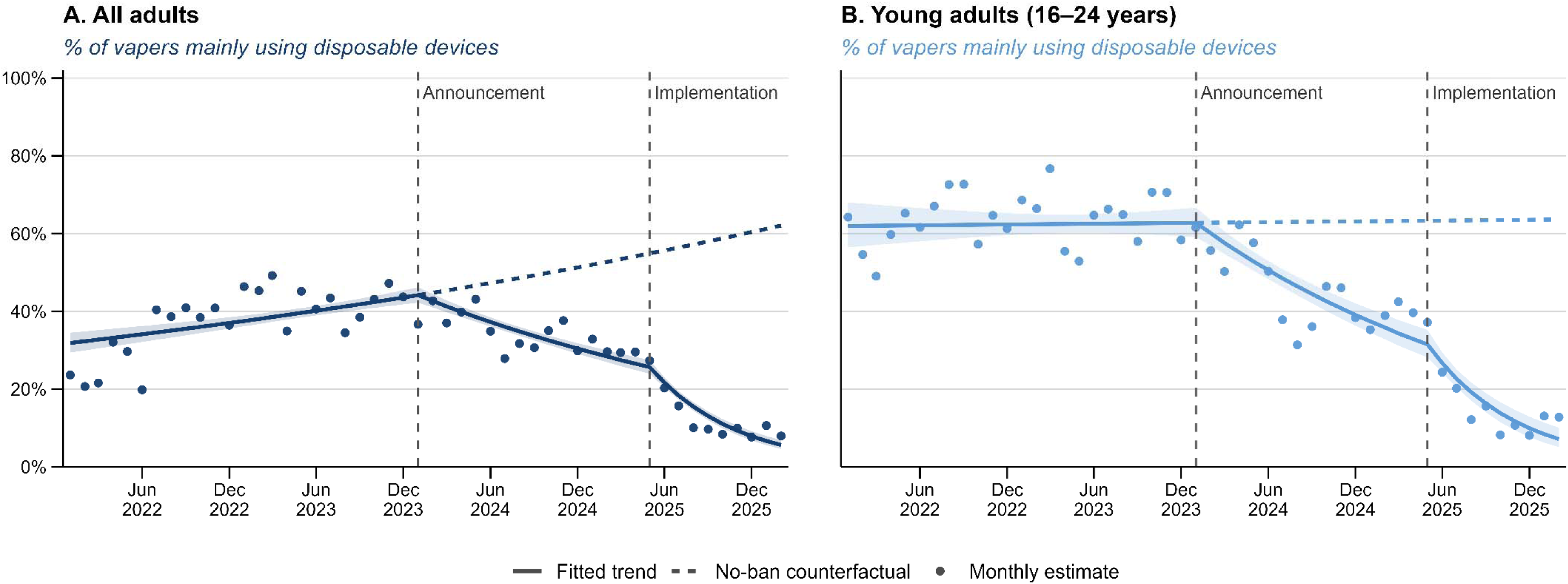
Changes in the proportion of adult (N=11,115) and young adult (N=2,473) vapers who mainly use disposable devices in Great Britain following the announcement (January 2024) and implementation (June 2025) of the ban. Solid lines show fitted trends from seasonality-adjusted segmented regression models (generalised additive models with a log link). Dotted lines show estimates under a “no-ban” counterfactual, assuming pre-announcement trends continued unchanged. Points show raw monthly estimates. All estimates are weighted to be representative of the Great Britain population.

Similar changes were observed among young adult vapers (aged 16–24 years), but from a higher starting point (**Table 1**; **Figure 1B**). Before the announcement, the proportion of young adult vapers mainly using disposables was approximately stable (RR_trend_=1.01, 95% CI=0.94–1.07), at around 62% (**Table S2**). After the announcement, the trend shifted sharply downward (RR_Δtrend_=0.59, 95% CI=0.51–0.69), with prevalence declining thereafter (post-announcement RR_trend_ =0.60, 95% CI=0.54–0.66). The decline accelerated following implementation (RR_Δtrend_=0.23, 95% CI=0.13–0.41), producing a very steep post-implementation decline (RR_trend_=0.14, 95% CI=0.08–0.23). By February 2026, 7.1% (95% CI=5.1–10.1) of young adult vapers mainly used disposables, versus 63.6% (52.7–76.7) under the no-ban counterfactual scenario, a difference of -56.5 percentage points (-68.5 to -44.4).

Descriptive data (**Figure S1**) show that the decline in vapers reporting mainly using disposable devices between 2024 and 2026 was mostly offset by an increase in the proportion using reusable pre-filled cartridge devices (from around one in ten vapers to one in three). Refillable tank devices were the most popular type used in all but one of the quarters, and the proportion who did not know what type of device they mainly used remained low (<5%) throughout.

### Vaping prevalence

Trends in current vaping prevalence also changed after the announcement and, to a lesser extent, after implementation of the ban (**Table 1**; **Figure 2A-B**). In adults, current vaping had been increasing before the announcement (RR_trend_=1.20, 95% CI=1.17–1.25). This increase slowed after announcement (RR_Δtrend_=0.91, 95% CI=0.85–0.97), although prevalence still increased modestly during the post-announcement period (RR_trend_=1.09, 95% CI=1.04–1.14). After implementation there was a further downward change in trend (RR_Δtrend_=0.86, 95% CI=0.75–0.98), such that the post-implementation trend was broadly flat (RR_trend_=0.94, 95% CI=0.84–1.04). By February 2026, observed current vaping prevalence among adults was 13.6% (95% CI=12.8–14.4), compared with 18.8% (17.0–20.7) under the no-ban counterfactual, a difference of -5.2 percentage points (-7.1 to -3.3).

**Figure 2.**
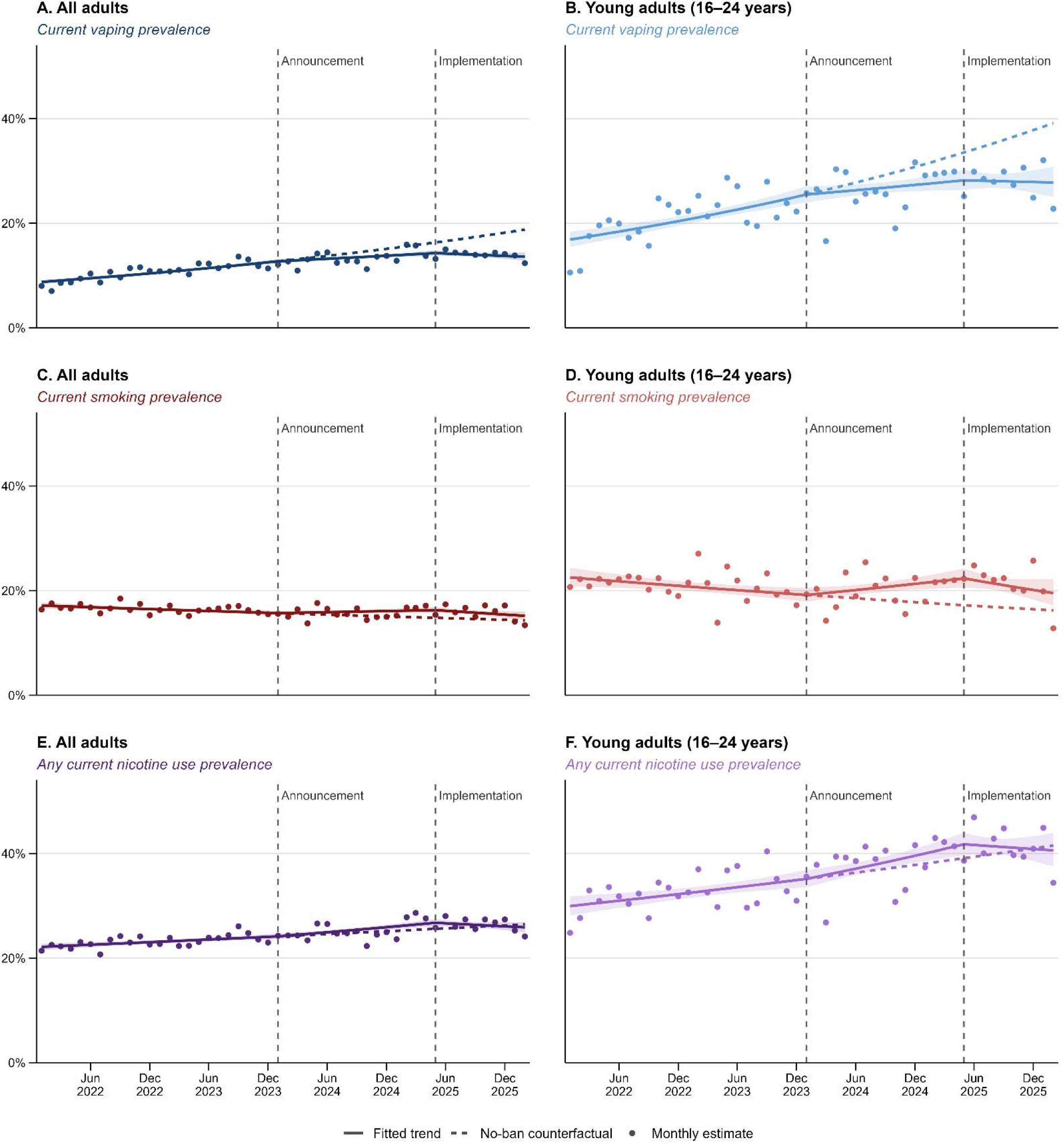
Changes in vaping, smoking, and any nicotine use among adults (N=118,946) and young adults (N=12,042) in Great Britain following the announcement (January 2024) and implementation (June 2025) of the ban. Solid lines show fitted trends from seasonality-adjusted segmented regression models (generalised additive models with a log link). Dotted lines show estimates under a “no-ban” counterfactual, assuming pre-announcement trends continued unchanged. Points show raw monthly estimates. All estimates are weighted to be representative of the Great Britain population.

Among young adults, current vaping prevalence was increasing before the announcement (RR_trend_=1.23, 95% CI=1.16–1.31), with evidence of a downward change in trend afterwards (RR_Δtrend_ =0.88, 95% CI=0.78–0.99). Prevalence increased more slowly during the post-announcement period (RR_trend_=1.08, 95% CI=1.00–1.17). The evidence of further change after implementation was unclear (RR_Δtrend_=0.91, 95% CI=0.71–1.16), leaving a post-implementation trend broadly stable (RR_trend_=0.98, 95% CI=0.81–1.18). By February 2026, current vaping prevalence among young adults was 27.8% (95% CI=25.0–30.8), versus 39.1% (32.8–46.7) in the no-ban counterfactual scenario, a difference of -11.3 percentage points (-18.6 to -4.1).

### Smoking prevalence

Among all respondents, trends in current smoking prevalence changed after the announcement and again after implementation (**Table 1**; **Figure 2C-D**). In adults, current smoking had been slowly declining before the announcement (RR_trend_=0.96, 95% CI=0.93–0.98). This trend changed after February 2024 (RR_Δtrend_=1.07, 95% CI=1.01–1.14), such that smoking prevalence was broadly flat to slightly increasing during the post-announcement period (RR_trend_=1.03, 95% CI=0.99–1.07). After implementation there was an uncertain downward change in trend (RR_Δtrend_=0.89, 95% CI=0.79–1.00), such that the post-implementation trend appeared to be downward (RR_trend_=0.91, 95% CI=0.83–1.00). By February 2026, observed current smoking prevalence among adults was 15.2% (95% CI=14.4–16.0), compared with 14.3% (13.2–15.5) under the no-ban counterfactual, an uncertain difference of +0.9 percentage points (-0.5 to +2.2).

Among young adults, current smoking prevalence had been slowly declining before the announcement (RR_trend_=0.92, 95% CI=0.87–0.98), but there was evidence of an upward change in trend after February 2024 (RR_Δtrend_=1.22, 95% CI=1.06–1.40). Smoking prevalence then increased during the post-announcement period (RR_trend_=1.12, 95% CI=1.02–1.23). After implementation there was an uncertain downward change in trend (RR_Δtrend_=0.75, 95% CI=0.56–1.00), leaving the post-implementation trend consistent with a decline rather than continued increase (RR_trend_=0.84, 95% CI=0.66–1.05). By February 2026, current smoking prevalence among young adults was 19.5% (95% CI=17.2–22.2), versus 16.2% (13.4–19.6) in the no-ban counterfactual scenario, an uncertain difference of +3.3 percentage points (-0.5 to +7.1).

In unplanned analyses separating daily from non-daily cigarette smoking, the patterns were uncertain but suggested that changes in current smoking may have been driven more by non-daily than daily smoking (**Table S3**; **Figure S2**). Daily smoking declined before the announcement and was broadly flat thereafter, with little evidence of a further change after implementation. By contrast, non-daily smoking was broadly stable before the announcement, increased during the post-announcement period, and then possibly declined after implementation, although end-of-series differences from the no-ban counterfactual were uncertain in both adults and young adults.

### Nicotine use prevalence

Among all respondents, trends in any current nicotine use changed little after the announcement but shifted downward after implementation (**Table 1**; **Figure 2E-F**). In both adults and young adults, prevalence had been slowly increasing before the announcement, but there was some uncertain evidence of a small accelerated increase following the announcement (adults: RR_Δtrend_=1.03, 95% CI=0.99–1.08; young adults: RR_Δtrend_=1.05, 95% CI=0.96–1.15). After implementation, however, there was a downward change in trend in adults (RR_Δtrend_=0.89, 95% CI=0.81–0.97) and some uncertain evidence of a downward change in young adults (RR_Δtrend_=0.85, 95% CI=0.70–1.02), leaving post-implementation trends close to stable in both groups (adults: RR_trend_=0.96, 95% CI=0.89–1.03; young adults: RR_trend_=0.96, 95% CI=0.84–1.11). By February 2026, the end result was that observed prevalence was similar to the steadily increasing no-ban counterfactual in all adults [25.9% vs 26.4%; difference=-0.6 percentage points, 95% CI=-2.4 to +1.3] and young adults [40.6% vs 41.5%; difference=-0.9 percentage points, 95% CI=-7.0 to +5.2]; however if the post implementation stability persists, then it would soon diverge and be less than the steadily increasing counterfactual.

## Discussion

In this study using data from a representative sample of adults in Great Britain, disposable vape use started falling after the January 2024 announcement of a forthcoming disposable vape ban, and then fell much more steeply after the ban was implemented in June 2025. By February 2026, 6% of all (≥16 years) and 7% of young adult (16–24 years) vapers reported mainly using disposable devices, compared with 62% and 64%, respectively, in a no-ban counterfactual scenario in which pre-announcement trends were assumed to continue unchanged. Current vaping prevalence was also lower than this no-ban counterfactual in both all (-5 percentage points) and young adults (-11 percentage points), suggesting that the ban was associated with some reduction in vaping overall, in addition to a shift away from disposables. Smoking prevalence had been falling before the announcement, but this decline stopped in adults and reversed into an increase in young adults after the announcement; after implementation, smoking trends shifted downward again in both groups.

These results show that behaviour may have changed in anticipation of the policy, rather than only once legal restrictions came into force. This is consistent with previous evidence on other tobacco and nicotine policies where announcement effects have preceded implementation.[26,27] Several mechanisms may explain this pattern. First, the announcement may have increased awareness of the environmental impact of disposable vapes or signalled official concern about these products, prompting some people to switch to rechargeable devices, stop vaping, or revert to smoking.[14,28] Second, manufacturers rapidly adapted to the impending ban by introducing rechargeable products designed to closely resemble popular disposable models. These “disposable-like” products match disposables in appearance, flavours, branding, price, and ease of use, differing primarily in the inclusion of a charging port and replaceable cartridges.[29] If these shared features — rather than disposability itself — were the main drivers of appeal, as some evidence suggests,[30–32] such products may have enabled a straightforward switch in anticipation of the ban.

Implementation of the ban was associated with a further and much steeper decline in the proportion of vapers mainly using disposable devices, showing that restricting legal sale and supply had effects beyond those seen after announcement alone. Even so, more than one in 20 vapers still reported mainly using disposables eight months after implementation. This may reflect consumers bulk-buying ahead of the ban, cross-border purchasing or continued illegal sales (as suggested by media reports[12]), but it may also reflect that some users continued to classify “disposable-like” rechargeable devices as disposables, and may have continued to treat them as such.

Increases in vaping prevalence slowed following the ban’s announcement and plateaued following implementation, such that, by February 2026, it was lower than expected under the no-ban counterfactual, especially among young adults. This pattern is consistent with the announcement and implementation of the ban reducing uptake of vaping and/or increasing cessation of vaping. It is also possible that vaping prevalence would have slowed or plateaued in the absence of regulation, as the earlier rapid growth in vaping approached a natural ceiling.[33] However, the absence of a similarly clear reduction in overall nicotine use suggests that most people substituted towards other nicotine products rather than stopping nicotine use altogether.

Smoking trends were more nuanced. The interruption in the prior downward trend after the announcement, and the increase observed among young adults, could reflect some individuals responding by smoking instead, potentially because of anticipated reduced access to preferred vaping products or changes in perceptions of vaping following the policy announcement.[34] Qualitative studies have identified this as a potential reaction among some users.[14] Unplanned analyses suggested that these changes may have been driven more by non-daily than daily smoking, with daily smoking continuing to decline or remaining broadly stable throughout. While these exploratory findings were imprecise and should be interpreted cautiously, they may indicate experimentation or intermittent smoking rather than substantial increases in daily smoking. After implementation, smoking trends appeared to shift downward again, but the reasons for this are unclear. These changes were relatively small, estimates were imprecise, and they may also reflect other contemporaneous influences rather than effects of the ban itself.[35] Nevertheless, smoking is lethal and exposes users (as well as bystanders) to a wider range and substantially higher levels of toxicants and carcinogens than vaping; even non-daily smoking carries considerable risks.[36,37] Any increase in smoking following vaping regulation could therefore easily offset public health gains from reductions in vaping.[38] Evidence of substitution between products following tightened vaping regulation[39–42] underscores the importance of proportionate measures and careful evaluation of vaping policy.

The prevalence of nicotine use has not clearly declined among young adults, and smoking is only decreasing slowly, if at all. This highlights the need for further measures to reduce uptake and support cessation. The UK government’s recent increased investment in stop smoking services, mass media campaigns, and the Swap to Stop scheme may help accelerate declines in smoking.[25] In addition, the Tobacco and Vapes Act 2026 will introduce stricter regulations, including the “smoke-free generation” policy, which will prohibit the sale of combustible tobacco products to people born on or after 1^st^ January 2009, as well as bans on advertising, sponsorship, free distribution, and vending machine sales for all nicotine products. The Act also provides powers to regulate vape product design, packaging, point-of-sale displays, and flavours; to extend smokefree legislation to prohibit vaping in some public places; and to create a retail licensing scheme for nicotine products. The detailed provisions of these restrictions will be set out through secondary legislation and public consultation.[43] Careful use of these powers may be needed given the emergence of rechargeable “disposable-like” vape products that retain many features that made disposables appealing, and because these powers also extend to oral nicotine pouches, which have seen a sharp increase in use among young men over the past five years.[18,29]

This study has several strengths. It used nationally-representative monthly data and was able to examine changes following both the announcement and implementation of the ban. The relatively long pre-implementation period helped to characterise underlying trends, and the monthly data provided greater temporal granularity and timeliness than annual surveys. However, several limitations should be considered. First, only nine months of post-implementation data were available, so estimates for some outcomes were imprecise. Second, as with all interrupted time-series analyses, the results cannot definitively establish causality, and other contemporaneous changes may also have influenced behaviour. Third, while the modelled no-ban counterfactual provides a useful reference for interpreting the magnitude of departures from pre-announcement trends, it should not be treated as an observed alternative history. For these contrasts to reflect a causal effect of the ban, it must be assumed that pre-announcement trends would have continued unchanged in the absence of the ban; this assumption is untestable and may be questioned, particularly for outcomes that were changing rapidly before the announcement. For example, trends may departed from linearity for other reasons unrelated to the ban, such as vaping prevalence approaching a natural plateau. Finally, the repeat cross-sectional design does not allow assessment of changes within individuals, so we cannot disentangle the extent to which observed changes in the prevalence of vaping, smoking, or nicotine use reflect new uptake, switching between products, or cessation.

The disposable vape ban in Great Britain was associated with substantial changes following both announcement and implementation, including a marked reduction in disposable vape use and a plateau in the previous rise in vaping prevalence. However, declines in smoking slowed (and reversed among young adults) after the announcement and only resumed gradually after implementation. Persistently high levels of vaping among young adults, alongside slow declines in smoking, suggest that further policy measures may be needed. Measures in the Tobacco and Vapes Act 2026 may help reduce the appeal and accessibility of nicotine and tobacco products to young people. As secondary legislation on vaping products is developed, its effects on both vaping and smoking should be carefully considered and evaluated.

## Data Availability

Monthly data and analysis code were provided openly to all through the Open Science Framework (https://osf.io/beac8).

## Ethics statements

Participants provided informed consent, and the UCL Ethics Committee gave ethical approval for the Smoking Toolkit Study (ID 0498/001). The data were not collected by UCL and were received in anonymised form.

## Data sharing statement

Monthly data and analysis code were provided openly to all through the Open Science Framework (https://osf.io/beac8). HTB and VB have directly accessed and verified the underlying data reported in the manuscript.

## Acknowledgements/Funding

The Smoking Toolkit Study is supported by Cancer Research UK (PRCRPG-Nov21\100002) and the Behavioural Research UK Leadership Hub, which is funded by the Economic and Social Research Council (ES/Y001044/1). Cancer Research UK provides salary support to HTB, SC, VB, and SJ, while VB is additionally supported by the Behavioural Research UK Leadership Hub. To support open access, the author has applied a CC BY licence to any Author Accepted Manuscript resulting from this submission. The funding bodies had no involvement in the study design, data collection, analysis, interpretation, manuscript preparation, or the decision to submit.

## Transparency

The authors, the manuscript’s guarantors, affirm that this manuscript is an honest, accurate, and transparent account of the study being reported; that no important aspects of the study have been omitted; and that any discrepancies from the study as originally planned and registered have been explained.

## Contributors

HTB led the analysis and drafting of the manuscript. All authors contributed to the development of the research questions, interpretation of findings, manuscript revision. All authors approved the final version of the manuscript.

