## Supplementary Material for "Impact of the disposable vape ban in Great Britain: a representative interrupted time-series study 2022-2026"

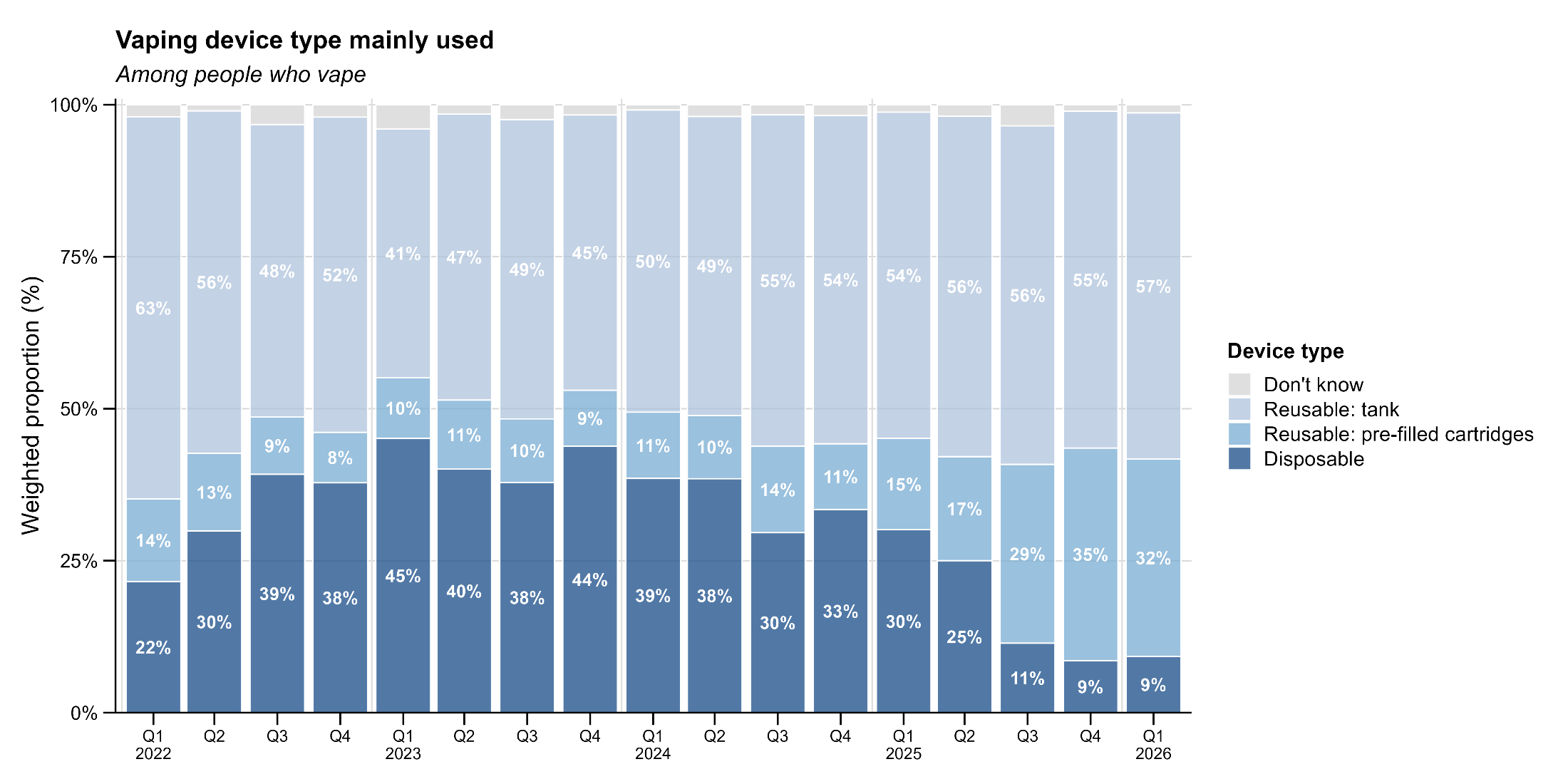

**Figure S1. Changes in the distribution of device types among adult (N=11,332) vapers in Great Britain from January 2022 to February 2026.** Stacked bars show the weighted proportion of vapers reporting each main device type, including disposable devices, reusable devices with pre-filled cartridges, reusable tank systems (including mods), and unknown device type. Percentages are shown for categories contributing at least 5% of the total. All estimates are weighted to be representative of the Great Britain population.

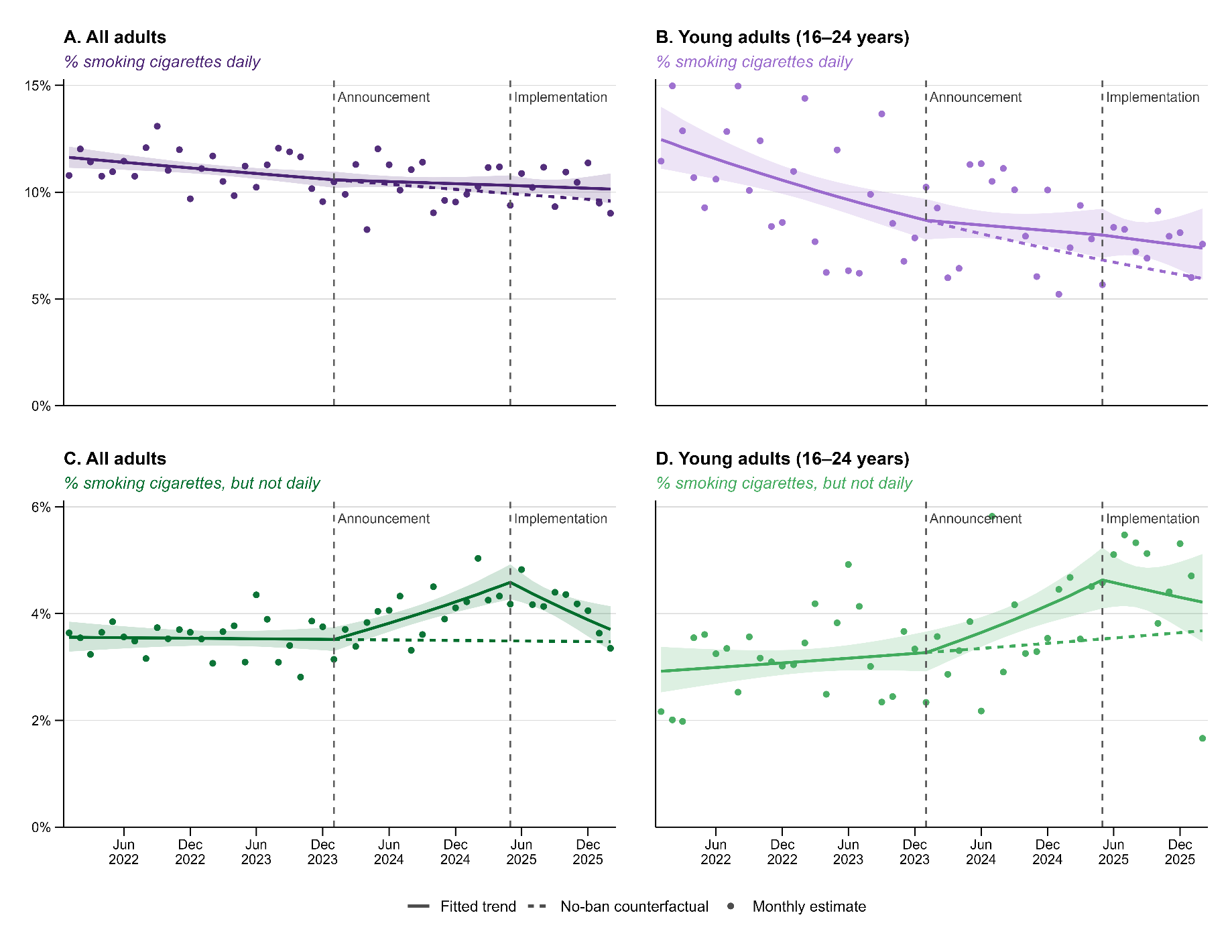

**Figure S2. Changes in the prevalence of daily and non-daily cigarette smoking among adults (N=118,946) and young adults (N=12,042) in Great Britain following the announcement (January 2024) and implementation (June 2025) of the ban.** Solid lines show fitted trends from seasonality-adjusted segmented regression models (generalised additive models with a log link). Dotted lines show estimates under a “no-ban” counterfactual, assuming pre-announcement trends continued unchanged. Points show raw monthly estimates. All estimates are weighted to be representative of the Great Britain population.

**Table S1. Sociodemographic and nicotine use characteristics (aged ≥16, N=118,946) of sample surveyed in Great Britain between January 2022 and February 2026.**

|  |  | Pre-announcement  (Jan 22 to Jan 24) | |  | Post-announcement  (Feb 24 to May 25) | |  | Post-implementation  (Jun 25 to Feb 26) | |
| --- | --- | --- | --- | --- | --- | --- | --- | --- | --- |
|  |  | n | % |  | n | % |  | n | % |
| **Overall** |  | 59,089 |  |  | 38286 |  |  | 21571 |  |
| **Country** | England | 41606 | 86.4 |  | 26999 | 86.4 |  | 15378 | 86.4 |
|  | Wales | 6224 | 4.9 |  | 4106 | 5.0 |  | 2258 | 4.9 |
|  | Scotland | 11259 | 8.7 |  | 7181 | 8.7 |  | 3935 | 8.7 |
| **Age (years)** | 16-17 | 847 | 1.8 |  | 387 | 1.3 |  | 175 | 1.0 |
|  | 18-24 | 5523 | 11.5 |  | 3331 | 11.6 |  | 1779 | 11.8 |
|  | 25-34 | 8013 | 16.6 |  | 5221 | 16.7 |  | 2688 | 16.8 |
|  | 35-44 | 7986 | 15.6 |  | 5055 | 15.8 |  | 2673 | 15.8 |
|  | 45-64 | 20089 | 31.6 |  | 12386 | 31.7 |  | 6852 | 31.7 |
|  | 65+ | 16598 | 22.8 |  | 11892 | 22.9 |  | 7400 | 22.8 |
|  | Refused | 33 | 0.1 |  | 14 | 0.0 |  | 4 | 0.0 |
| **Gender** | Men | 29159 | 48.4 |  | 19111 | 48.3 |  | 11065 | 48.2 |
|  | Woman | 29219 | 50.4 |  | 18622 | 50.3 |  | 10161 | 50.2 |
|  | In another way | 711 | 1.2 |  | 553 | 1.4 |  | 345 | 1.6 |
| **Social grade** | ABC1 (more advantaged) | 39614 | 56.0 |  | 26410 | 56.0 |  | 14999 | 56.2 |
|  | C2DE (less advantaged) | 19475 | 44.0 |  | 11876 | 44.0 |  | 6572 | 43.8 |
| n is unweighted. Percentages are weighted to be representative of the adult (≥16 years) population in Great Britain. | | | | | | | | | |

**Table S2. Seasonally-adjusted prevalence estimates at key time points surrounding the announcement (January 2022) and implementation (June 2025) of the disposable vape ban in Great Britain.**

|  |  | Among current vapersᵃ |  | Among all adultsᵇ | | |
| --- | --- | --- | --- | --- | --- | --- |
|  |  | Mainly use disposables |  | Current vaping | Current smoking | Any current nicotine use |
| **All adults, % [95% CI]** | |  |  |  |  |  |
|  | Start of series (Jan 2022) | 31.9 [29.4–34.5] |  | 8.8 [8.4–9.2] | 17.2 [16.6–17.7] | 22.1 [21.5–22.8] |
|  | Month before announcement (Jan 2024) | 44.1 [42.2–46.2] |  | 12.7 [12.3–13.2] | 15.7 [15.3–16.2] | 24.2 [23.6–24.7] |
|  | Month before implementation (May 2025) | 25.6 [23.9–27.5] |  | 14.3 [13.8–14.8] | 16.3 [15.7–16.9] | 26.7 [26.1–27.4] |
|  | End of series (Feb 2026) | 5.6 [4.6–6.9] |  | 13.6 [12.8–14.4] | 15.2 [14.4–16.0] | 25.9 [24.9–26.9] |
| **Young adults (16–24 years), % [95% CI]** | |  |  |  |  |  |
|  | Start of series (Jan 2022) | 62.0 [56.4–68.0] |  | 16.9 [15.5–18.4] | 22.5 [20.8–24.4] | 29.9 [28.2–31.8] |
|  | Month before announcement (Jan 2024) | 62.8 [59.1–66.7] |  | 25.5 [24.0–27.1] | 19.2 [17.9–20.5] | 35.1 [33.5–36.8] |
|  | Month before implementation (May 2025) | 31.5 [28.1–35.2] |  | 28.2 [26.3–30.2] | 22.4 [20.6–24.2] | 41.8 [39.7–44.0] |
|  | End of series (Feb 2026) | 7.1 [5.1–10.1] |  | 27.8 [25.0–30.8] | 19.5 [17.2–22.2] | 40.6 [37.5–43.9] |
| **Abbreviations:** RR, risk ratio; CI, confidence interval. **Notes:** Results are derived from segmented regression analyses using generalised additive models with a log link, adjusted for seasonality. Coefficients for trends are shown in Table 1, and estimates at every timepoint are provided in Figures 1 and 2.  ᵃ Among respondents who reported current vaping and provided information on main device type used (N=11,115 adults; N=2,473 young adults aged 16–24).  ^b^ Among all respondents who provided their smoking and vaping status (N=118,946 adults; N=12,042 young adults aged 16–24).  ^c^ Among respondents who had smoked and attempted to quit smoking in the past year (N=6,634 adults; N=1,343 young adults aged 16–24). | | | | | | |

**Table S3. Trends in daily and non-daily smoking prevalence before and after announcement (January 2022) and implementation (June 2025) of the disposable vape ban in Great Britain.**

|  |  |  | **Among all adults^a^** | |
| --- | --- | --- | --- | --- |
|  |  |  | **Daily smoking** | **Non-daily smoking** |
| **All adults** | |  |  |  |
|  | **Annual trends, RR [95% CI]** |  |  |  |
|  | Pre-announcement trend |  | 0.95 [0.92–0.99] | 0.99 [0.94–1.05] |
|  | *Change in trend after announcement* |  | 1.03 [0.96–1.11] | 1.23 [1.08–1.39] |
|  | Post-announcement trend |  | 0.98 [0.93–1.03] | 1.22 [1.13–1.32] |
|  | *Change in trend after implementation* |  | 1.00 [0.85–1.17] | 0.62 [0.48–0.79] |
|  | Post-implementation trend |  | 0.98 [0.86–1.11] | 0.75 [0.62–0.92] |
|  | **End-of-series prevalence (Feb 2026), % [95% CI]** |  |  |  |
|  | Observed |  | 10.1 [9.5–10.9] | 3.7 [3.3–4.1] |
|  | No-ban counterfactual scenario |  | 9.6 [8.7–10.6] | 3.5 [2.9–4.2] |
|  | *Difference* |  | 0.6 [-0.6 to 1.7] | 0.2 [-0.5 to 0.9] |
| **Young adults (16–24 years)** | |  |  |  |
|  | **Annual trends, RR [95% CI]** |  |  |  |
|  | Pre-announcement trend |  | 0.83 [0.76–0.92] | 1.06 [0.95–1.18] |
|  | *Change in trend after announcement* |  | 1.13 [0.90–1.41] | 1.23 [0.98–1.54] |
|  | Post-announcement trend |  | 0.94 [0.81–1.09] | 1.30 [1.13–1.50] |
|  | *Change in trend after implementation* |  | 0.96 [0.58–1.59] | 0.68 [0.43–1.07] |
|  | Post-implementation trend |  | 0.90 [0.60–1.34] | 0.88 [0.62–1.25] |
|  | **End-of-series prevalence (Feb 2026), % [95% CI]**^b^ |  |  |  |
|  | Observed |  | 7.4 [5.9–9.2] | 10.5 [8.7–12.8] |
|  | No-ban counterfactual scenario |  | 6.0 [4.4–8.0] | 9.2 [6.7–12.7] |
|  | *Difference* |  | 1.4 [-0.9 to 3.7] | 1.3 [-2.1 to 4.8] |
| **Abbreviations:** RR, risk ratio; CI, confidence interval. **Notes:** Results are derived from survey-weighted segmented regression analyses using generalised additive models with a log link, adjusted for seasonality.  ᵃ Among all respondents who provided their smoking and vaping status (N=118,946 adults; N=12,042 young adults aged 16–24).  ^b^ No-ban estimates represent model-based predictions at February 2026 obtained by extrapolating the pre-announcement trend forward. The observed–no ban difference is calculated as the modelled observed prevalence minus the extrapolated no-ban prevalence at February 2026. | | | | |
